# Effectiveness of BNT162b2 and ChAdOx1 against SARS-CoV-2 household transmission: a prospective cohort study in England

**DOI:** 10.1101/2021.11.24.21266401

**Authors:** Samuel Clifford, Pauline Waight, Jada Hackman, Stephane Hué, Charlotte M Gower, Freja CM Kirsebom, Catriona Skarnes, Louise Letley, Jamie Lopez Bernal, Nick Andrews, Stefan Flasche, Elizabeth Miller

## Abstract

**Background:** The ability of SARS-CoV-2 vaccines to protect against infection and onward transmission determines whether immunisation can control global circulation. We estimated effectiveness of BNT162b2 and ChAdOx1 vaccines against acquisition and transmission of the Alpha and Delta variants in a prospective household study in England.

**Methods:** Adult index cases in the community and their household contacts took oral-nasal swabs on days 1, 3 and 7 after enrolment. Swabs were tested by RT-qPCR with genomic sequencing conducted on a subset. We used Bayesian logistic regression to infer vaccine effectiveness against acquisition and transmission, adjusted for age, vaccination history and variant.

**Findings:** Between 2 February 2021 and 10 September 2021 213 index cases and 312 contacts were followed up. After excluding households lacking genomic proximity (N=2) or with unlikely serial intervals (N=16), 195 households with 278 contacts remained of whom 113 (41%) became PCR positive. Delta lineages had 1.64 times the risk (95% Credible Interval: 1.15 – 2.44) of transmission than Alpha; contacts older than 18 years were 1.19 times (1.04 - 1.52) more likely to acquire infection than children. Effectiveness of two doses of BNT162b2 against transmission of Delta was 31% (−3%, 61%) and 42% (14%, 69%) for ChAdOx1, similar to their effectiveness for Alpha. Protection against infection with Alpha was higher than for Delta, 71% (12%,95%) vs 24% (−2%, 64%) respectively for BNT162b2 and 26% (−39%, 73%) vs 14% (−5%, 46%) respectively for ChAdOx1.

**Interpretation:** BNT162b2 and ChAdOx1 reduce transmission of the Delta variant from breakthrough infections in the household setting though their protection against infection is low.

**Funding:** This study was funded by the UK Health Security Agency (formerly Public Health England) as part of the COVID-19 response.

## Introduction

The rapid development of safe and effective COVID-19 vaccines using both novel and traditional platforms, is an unprecedented scientific achievement. The United Kingdom was the first country to launch a national COVID-19 vaccination programme with the rollout of the Pfizer-BioNTech mRNA vaccine (BNT162b2) on 8th December 2020, followed shortly after by the Oxford AstraZeneca adenovirus vector vaccine (ChAdOx1). By September 2021, over 40% of the world’s population had received at least one dose of a COVID-19 vaccine, whether an mRNA, adenovirus vector, or inactivated whole virion vaccine.^1^ In most countries, vaccine deployment has been focussed on direct protection of those individuals at greatest risk of a severe outcome of SARS-CoV-2 infection including the elderly and those with co-morbidities. Health care workers and others who, if infected, pose a transmission risk to vulnerable individuals, have also been identified as a priority group for vaccination.

The primary outcome of the efficacy trials of the currently authorised COVID-19 vaccines was symptomatic laboratory confirmed SARS-CoV-2 infection, with little information generated on protection against severe COVID-19 infection nor on the ability of the vaccines to prevent onward transmission in those infected. There is now a growing body of evidence from observational studies showing high protection against severe COVID-19 from inactivated whole virion, mRNA, and adenovirus vector vaccines but information on protection against transmission is still limited.^2^ Attempts have been made to infer protection against transmission by comparing the viral load in the nasopharynx of vaccinated individuals with breakthrough infections with that in unvaccinated cases, using Ct values as a proxy.^3^ Other approaches have used routine diagnostic PCR testing data, constructing households based on individuals’ addresses or identifying them with contact tracing, and to estimate secondary attack rates by vaccination status of the index case. However, these studies are potentially subject to ascertainment bias as they are reliant on the testing behaviour of household contacts.^4–6^

Here we report the results of a prospective household transmission study set up by Public Health England (now the UK Health Security Agency) in January 2021 to assess the effect of the vaccination history of index cases with COVID-19 on transmission of SARS-CoV-2 to household contacts, and the protection afforded to vaccinated contacts under conditions of household exposure.

## Methods

### Data

#### Households

Details of household recruitment, ethics, data governance and laboratory testing has been reported elsewhere.^7^ In brief, infected index cases, identified via Pillar 2 community testing, and their consenting household contacts are recruited by study nurses, on average, 3 days after their initial PCR test. The vaccination status of index cases and their household contacts is obtained by data linkage with the National Immunisation Management System (NIMS) and checked with participants by the study nurse at the time of recruitment. Self-testing kits for the index case and household contacts to take combined nose and throat swabs on recruitment (Day 1), Day 3 and Day 7 are couriered to households and subsequently tested by dual target PCR at PHE Colindale (ORF and E genes). PCR positive swabs are sequenced as part of the COG-UK initiative.^8^

Household contacts were defined as infected if one or more swabs was PCR positive.

The household transmission study is ongoing and inclusion in this analysis is based on participants having returned at least one swab, being either unvaccinated or vaccinated with one or two doses of either BNT162b2 or ChAdOx1 with the vaccination dates available, and the age at time of recruitment and the date of onset of symptoms (fever, cough, runny nose, sore throat, shortness of breath, loss of taste or smell, nausea, diarrhea, muscle/body pain, headache or other) recorded.

### Statistical Analysis

All analysis was conducted in R 4.1.1^9^ with Bayesian models fit using the rjags package.^10^ The secondary attack rate (SAR) for each combination of case and contact is estimated here by predicting the probability an unseen contact acquires an infection from an infected case given the vaccination history and age of each. The effects of vaccination are presented in the results as risk ratios (RRs) for each vaccine product and number of doses compared to the unvaccinated group of the same age and household variant. The predicted SARs and RRs are summarised with medians and 95% credible intervals.

#### Household secondary attack rate

We fit a Bayesian hierarchical linear model with Bernoulli likelihood for the probability that a household contact of an index case acquires a SARS-CoV-2 infection within a week of recruitment. The model estimates both a protective effect for vaccinated contacts against infection and a reduction in transmission for vaccinated cases, which are assumed to be independent. The effect of the first dose is assumed to only occur 21 days after the vaccination is received, and an additional effect of the second dose requires at least 7 days have passed since the second vaccination as in the SIREN study which considers the effectiveness of BNT162b2 in healthcare workers in England^11^. These effects are assumed to depend on the vaccine product, and number of doses thereof, received by both the index case and the contact (Table A1). The probability of acquiring infection is also assumed to depend on the age of both the case and contact, and the circulating lineage. Vaccine efficacies are calculated as 1-relative risk in household secondary attack rates (SARs). For such the SARs were sampled during the MCMC sampling, for each combination of variant and case and contact vaccine status (1 or 2 doses for each product) and age group, against a baseline of that case-contact pair and variant in the absence of any vaccination.

#### Lineage

At the start of data collection, the B.1.1.7 (Alpha) SARS-CoV-2 variant was most prevalent in the United Kingdom, and an increasing proportion of swabs sequenced by Pillar 2 testing were identified as B.1.617.2 (Delta) variant over time^12^. Where sequencing was not available to determine the variant for a positive swab, the probability that it was the Delta variant was estimated from the date of sampling and a logistic regression model fit to the number of weekly cases identified through Pillar 2 that were either Alpha or Delta variant.

#### Participants’ age

Vaccine eligibility and type is correlated with age and date of vaccination. This is because from 7th April 2021 the BNT162b2 vaccine was recommended for under 30 years olds in preference to ChadOx1 with extension to 30-40 year olds from 7th May 2021^13^ and also because, apart from those in high risk groups, vaccination was not offered to the general 16-17 year old population until August 2021^14^ and the general 12-15 year old population until September 2021^15^. We account for age in the model by considering that children under 18 will have decreased susceptibility to infection, compared to adults,^16^ and that older adults are more likely to transmit.^17^ While the study did not specifically recruit only adult index cases, the minimum age of index cases was 21. The median age of index cases was 48 years and so we split adults into younger (18-49) and older (50+) age groups. Very few participants were older than 65 years so we do not distinguish between 50-64 and 65+ year olds. We did not adjust for prior infection status as information on this was incomplete at the time of data lock, nor for gender as this was previously shown not to be a factor in determining household transmission.^7^ Table A2 shows the age and vaccine status breakdown of index cases and their household contacts.

#### Infection history dynamics

PCR positivity relative to the onset of symptoms was estimated using data from all symptomatic cases and contacts, with pseudo-absences generated to simulate the time of infecting exposure. Comparison is made for each combination of vaccine product, number of doses, and variant against the corresponding unvaccinated group. Details of this modelling can be found in the Appendix.

#### Identification of non-household transmission

As per the study design, the index case for each household was by default considered to be the individual who presented for Pillar 2 testing. To reduce the risk of misclassification bias we excluded from the analyses all households where both the index case and an infected household contact were symptomatic and the index case’s symptoms appeared more than two days after the contact’s symptoms.

To further reduce the potential for misclassification bias, a phylogenetic approach was used to identify apparent secondary cases in the household who were in fact infected elsewhere. If none of the sequences from a contact clustered with at least one of the sequences from the household’s index case, then this was considered as evidence for an infection acquired outside of the household; therefore, the contact was excluded from the downstream analysis. Details of the phylogenetic approach can be found in the Appendix.

## Results

By September 10th, 2021, a total of 213 index cases and 312 contacts had been recruited and met the criteria for inclusion at that time. Two contacts were removed due to lack of genomic proximity (outlined below), which resulted in the removal of each of their households as there were no further contacts. The serial interval was 2 (95% range: -6 - 10) days. Sixteen households with their respective index cases and a total of 32 contacts were excluded from the main analysis because at least one infected household contact presented symptoms more than 2 days before the index case. Thus, the main analysis was performed on 195 index cases and their 278 contacts. Households had between 1 and 7 contacts, with a mean of 2.2, median of 2, and standard deviation of 1.2. The mode number of household contacts was 1.

Of the included individuals, 175 index cases (90%) and 113 (41%) contacts tested positive for SARS-CoV-2 at least once in the week since recruitment. Sequencing information was available for 122 (69%) and 81 (71%) of those, respectively.

Most (77%) index cases had received at least one dose of a vaccine at enrolment, whereas 52% of household contacts had (Table A1). 24% of contacts were less than 18 years old, and therefore not eligible for vaccination at the time. The proportion of at least partially vaccinated adult household contacts was 69%. Only 10 index cases (5%) were asymptomatic, reflecting the bias of Pillar 2 testing in the UK towards detecting mostly symptomatic infections.

### Prevalence of lineages

Of the 195 index cases analysed here, 99 were identified as infected with B.1.1.7 (Alpha), 24 with B.1.617.2 (Delta), 20 did not test positive again after recruitment, and 52 were of unknown lineage as their PCR-positive swabs had not yet been sequenced (Figure A3). Of the 72 individuals without information on the infecting lineage, we estimated that 18 were likely of Alpha and 54 were likely of Delta lineage based on the date of sampling (Figures A2, A3) and the national prevalence of lineages at the time. That is, 60% of index cases had an Alpha variant infection and the remainder were Delta.

### Identification of non-household transmission

Sequencing information for both index case and contact was available for 92 PCR positive case-contact pairs across 79 households. In total, 345 whole-genome sequences (including longitudinal samples) were available for analyses, a majority of which were of Alpha variant (82.6%) and the remainder were Delta (17.4%).

The phylogeny provided evidence that in two households the contact of the recruited index case had acquired infection elsewhere (Figure 1, households HH002 and HH007). Five households that did not form unique clusters in the phylogeny did not meet the exclusion criteria: in two a sequence from an index case did not cluster with the remaining household sequences but another sequence from the same index case did (HH004 and HH006) while the other three households did not have sufficient bootstrap support to be a part of a cluster (HH001, HH003, and HH005). Of the remaining households, 72 (91%), formed unique, household-specific clusters that included all and only sequences of members of the household, indicating likely direct transmission within the household.

**Figure 1:**
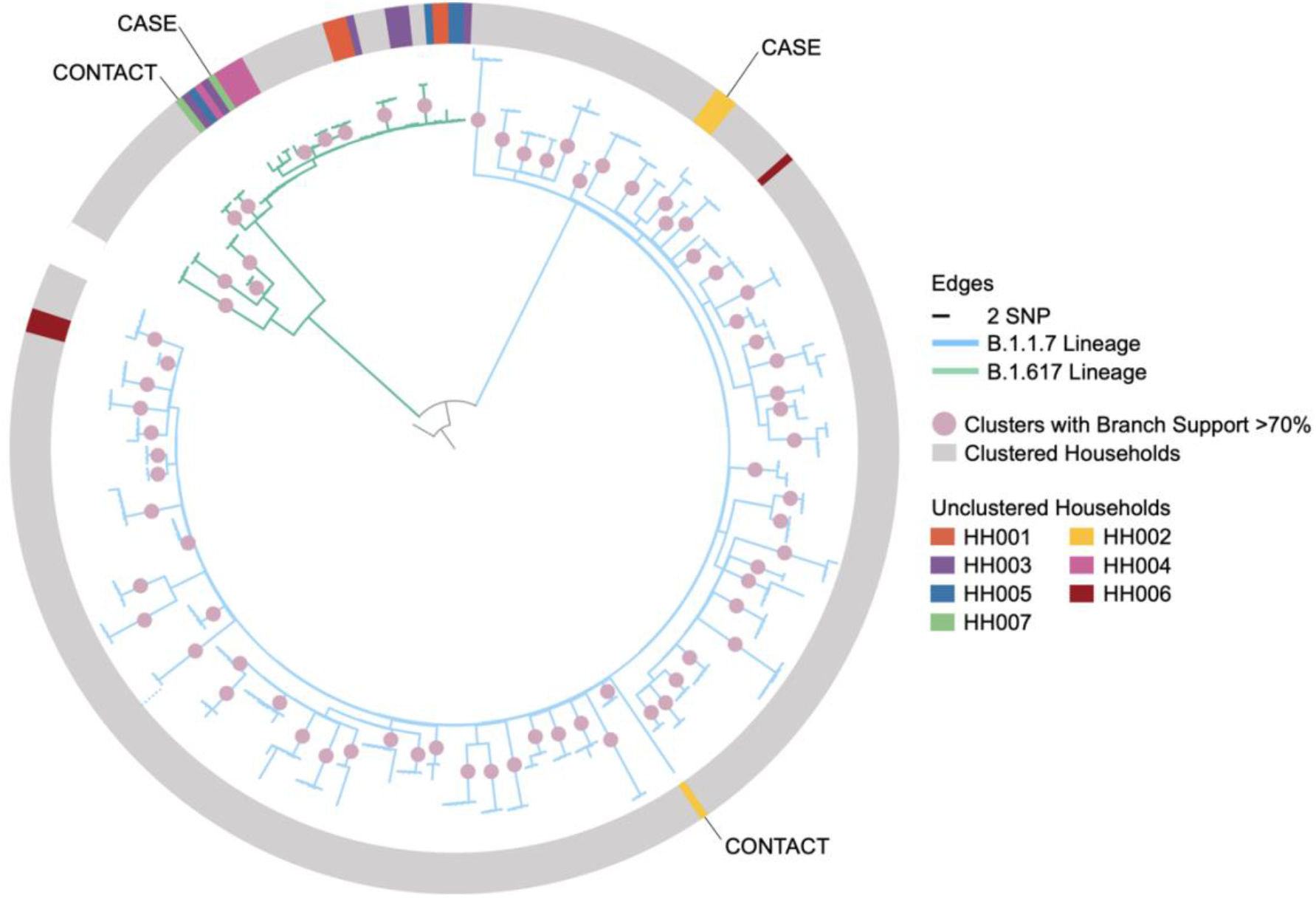
Maximum-likelihood phylogeny of household index cases and contacts’ sequences with 1,000 ultrafast bootstrap replicates rooted to the reference sequence with a scaled bar of 2 SNP (6.6 × 10^−5^ substitutions/site). The dotted line at bottom left indicates where a single long branch was collapsed for visualisation. The non-grey shading on the outer ring represents non-clustered households where sequences are coloured by their households. HH002 and HH007 were the only households where none of the contacts’ sequences clustered with their household’s index case’s and this is evidence the contact could have acquired the infection elsewhere and is thus excluded from the analysis.

### Age and lineage effects

We estimate that in the absence of vaccination of either case or contacts, Delta lineage infections were much more transmissible from non-elderly adult cases to adult contacts within the household than Alpha lineage infections (Risk Ratio: 1.64, 95% Credible Interval: 1.15, 2.44). Children younger than 18 years old were less likely as adults to acquire a Delta infection from non-elderly adults (RR: 0.84, 95%: 0.66, 0.96). Compared to a baseline of Delta index cases aged between 18 and 49, those 50 and over had 1.06 times the risk of transmitting their infection (95%: 1.00, 1.23).

### Effectiveness of vaccination

Either one or two doses of BNT162b2 provide a protective effect against infection from a symptomatic index case with Alpha variant SARS-CoV-2 with a vaccine effectiveness of 53%, (95% credible interval: 7%, 83%) and 71% (95% CrI: 12%, 95%), respectively (Table 1, Figure A4). At 4% (−21%, 44%) and 24% (−2%, 64%) the effectiveness of one and two doses of BNT162b2 against infection with the Delta variant was lower than against Alpha and was similar to the effectiveness offered by ChAdOx1 to either variant (Table A5) which, after two doses, had effectiveness against Alpha of 26% (−39%, 73%) and against Delta of 14% (−5%, 46%).

**Table 1:**
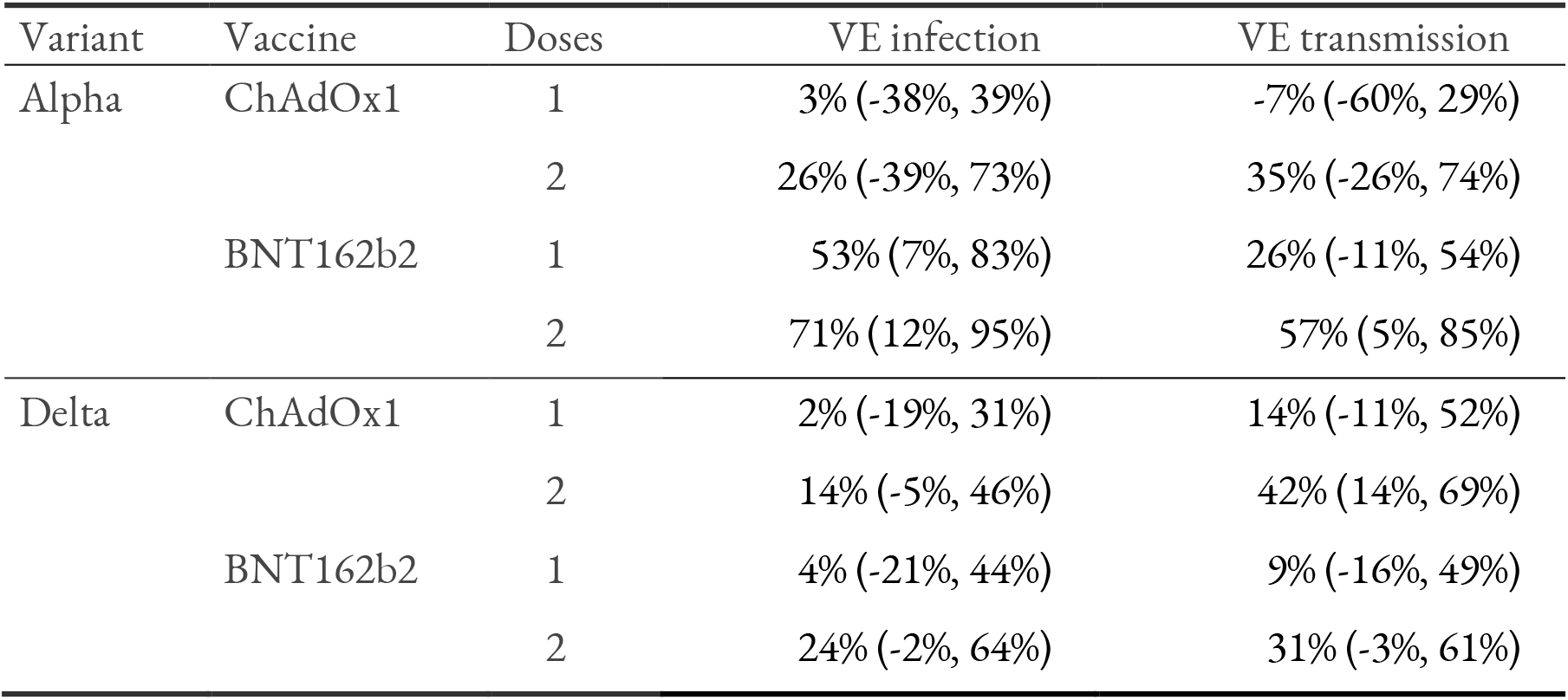
Median vaccine effectiveness (VE) and 95% credible intervals for infection protection in contacts and transmission reduction in cases, by variant, vaccine product, and number of doses.

We estimate that the effectiveness of one and two doses of BNT162b2 against onward transmission if infected with the Alpha variant was 26% (−11%, 54%) and 57% (5%, 85%) and for Delta variant one and two doses reduce transmission by 9% (−16%, 49%) and 37% (4%, 65%). RRs for the protective effect of BNT162b2 over ChAdOx1 for one and two doses of against both Alpha and Delta variants indicate that at 95% credibility there is no difference between the effectiveness of the two vaccine products (Table A5). Specifically, two doses of ChAdOx1 reduce transmission from an Alpha variant case by 35% (−26%, 74%) and from a Delta variant case by 42% (14%, 69%).

### Secondary attack rates

The estimated secondary household attack rate among adults in an unvaccinated household was 49% (34%, 63%) for the Alpha variant and 81% (57%, 96%) for the Delta variant (Figure 2). BNT162b2 is very effective against Alpha variant infection when either the case or contact are vaccinated, and especially when both have received two doses (Figure 2). SARs for Delta variant infection in unvaccinated case-contact pairs are substantially higher. Full (two dose) vaccination with either vaccine is still effective against Delta infection when both the case and contact are vaccinated, at least halving the SAR; e.g. case and contact both fully vaccinated with BNT162b2 has an SAR of 30% (13%, 57%). SARs for each combination of contact and case age, vaccine history and variant lineage are given in the appendix (Figures A5, A6). Notably, the reduced susceptibility to infection of (unvaccinated) under-18s results in SARs which are no greater than those seen in adult contacts who have received two doses of ChAdOx1.

**Figure 2:**
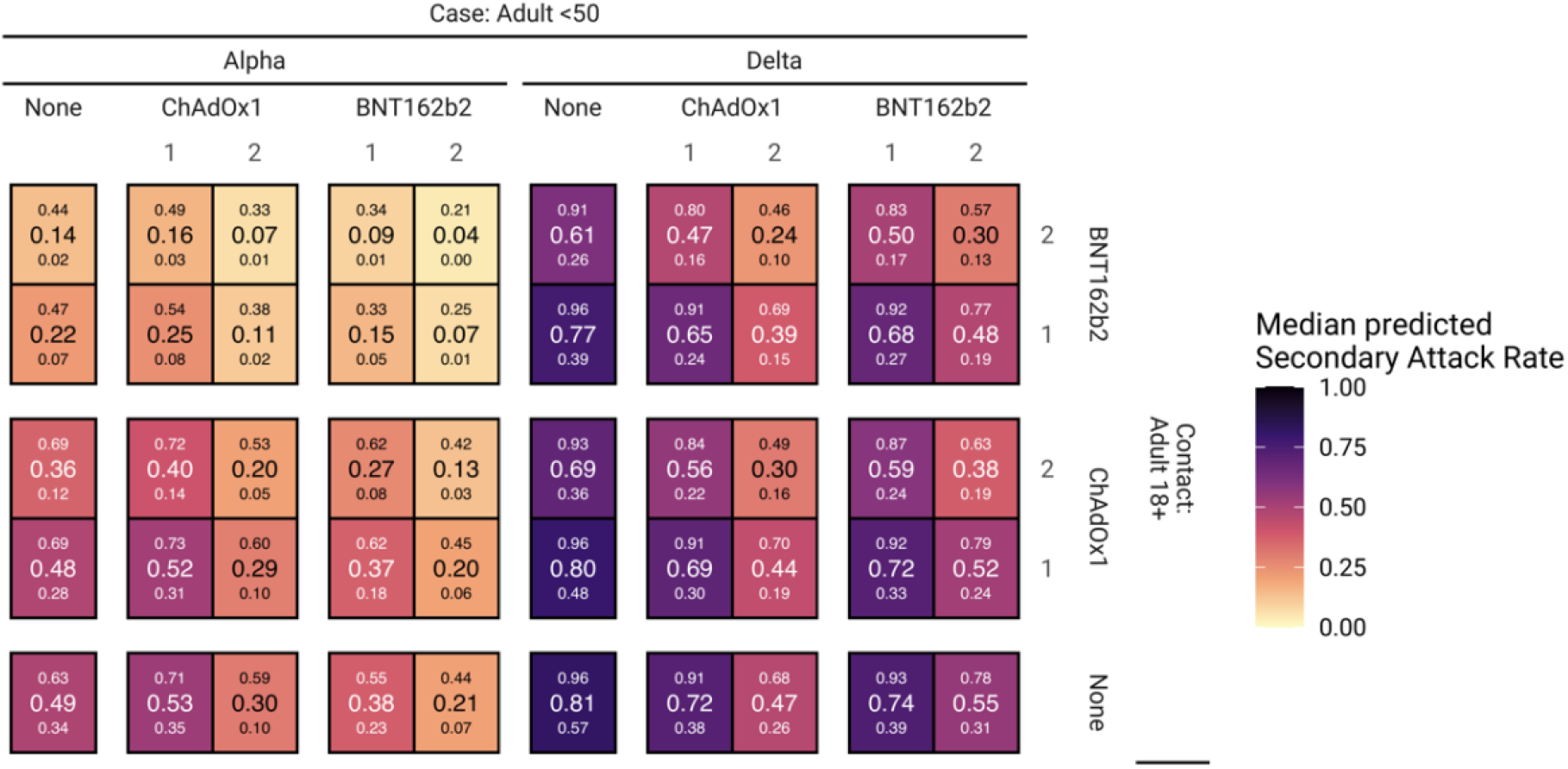
Predicted secondary attack rates (SARs) for each combination of vaccine status of case and contact. Large numbers inside cells are the median SAR, with the small numbers below and above corresponding to the 95% credible interval.

### Sensitivity analysis

Sensitivity analysis was conducted by including the 16 index case-contact pairs with serial intervals less than -2 days. This did not qualitatively change our results (Table A4). The absence of informative priors on the protective vaccine effects against infection led some of the vaccine effectiveness against infection in our study to be re-attributed to effectiveness against onward transmission or to age effects. Figure A4 provides a comparison of the exponentiated regression coefficients (odds ratios) for the vaccine effects for the main and sensitivity analyses as well as the informative prior used.

### Infection history dynamics

We estimate that within a week of symptom onset, the relative risk of testing PCR positive is near identical for vaccinated and unvaccinated participants. For cases infected with the Alpha variant, there was little difference in PCR positivity generally between vaccinated and unvaccinated cases while in cases infected with the Delta variant the proportion of participants with PCR detectable infection in participants fully vaccinated with BNT162b2 declined about 4 days before that in unvaccinated participants. At 2-3 days the effect in participants fully vaccinated with ChAdOx1 was slightly less pronounced.

## Discussion

In this prospective household-based study of SARS-CoV-2 infection we showed that both the ChAdOx1 and BNT162b2 vaccines are effective in reducing transmission of the Alpha and Delta variants from those who develop breakthrough infections despite having received two doses. The estimated vaccine effectiveness against acquisition of a Delta infection in the household setting was however low; 14% (−5%, 46%) and 24% (−2%, 64%) after two doses of ChAdOx1 and BNT162b2 respectively. This is lower than that estimated from cases presenting for Pillar 2 testing in the community for which the effectiveness of two doses of ChAdOx1 against symptomatic infection is estimated as 67.0% (61.3%, 71.8%) and 88.0% (85.3%, 90.1%) for BNT162b2.^18^ Effectiveness against acquisition of an Alpha infection in the household was substantially higher in our study than that against Delta but still lower than that estimated from Pillar 2 community testing. The lower protection against acquisition in the household likely reflects the prolonged and intense exposure that occurs in this setting. Similarly, although the effectiveness estimates against transmission were moderate at 42% (14%, 69%) and 31% (−3%, 61%) after 2 doses of ChadOx1 and BNT16b2 respectively, the protective effect in those with breakthrough infections may be higher in the community where exposure is less intense and of shorter duration. The reduction in duration of PCR positivity in breakthrough infections (average of 4 days shorter for the Delta variant for those infected after two doses of BNT162b2 and around 2-3 days for ChAdOx1) will also have more of an impact in the community than in the household setting where generation times between infections are short – around 3.5 days for the Delta variant.^19^ Our household contacts were actively followed up with repeated swabbing and showed the high secondary attack rates that occur in this setting; 81% for Delta infections in unvaccinated households but that reduced to 25-40% in households where both index case and contacts were fully vaccinated.

Our finding of a moderate level of protection against onward transmission from fully vaccinated individuals, with either vaccine and against either variant, is in apparent contrast to a study that similarly followed up contacts reported by the UK test and trace system prospectively, about 90% of whom were in the same household as the index case^20^. The study estimated a moderate effect of vaccination against infection but no difference in secondary attack rates with the delta variant between fully vaccinated and unvaccinated index cases (24% and 23% respectively). However, such estimates were neither controlled for age nor vaccination status of the contact. Interestingly only 4 out of 17 (24%) unvaccinated contacts were infected by fully vaccinated index cases, whereas 8 out of 20 (40%) unvaccinated contacts were infected by unvaccinated index cases; a reduction in transmission of 41% albeit based on very small numbers. Vaccine effectiveness against onward transmission of 40 to 80% has been suggested by several retrospective observational studies using either information on the household structure ^4^ or contact tracing ^5 6^ in combination with routine national COVID-19 notification systems to estimate reductions in secondary attack rates from breakthrough infections. While observational studies are prone to biases introduced by testing behaviour particularly for mild disease manifestations, our study combines prospectively collected data with a robust analytical framework to confirm that both vaccines reduce transmissibility of breakthrough infections in fully vaccinated individuals.

Among symptomatic index cases and contacts, we found a lower rate of PCR positivity within two weeks of symptom onset in all vaccinated groups (Figures 3 and A7). PCR positivity for Delta declined fastest (4 days ahead of unvaccinated) in individuals fully vaccinated with BNT162b2. These results largely mirror those in other studies that found enhanced clearance following vaccination,^20^ but raise the question whether enhanced clearance can be the driving mechanism for reduced transmission in a frequent contact household setting. Another mechanism may be that while positivity with the highly sensitive PCR test is similar to that in the unvaccinated, vaccination can reduce ^21^both peak viral load^3,22^ and viral shedding^23^, although such effects have not been reported in all studies and may be masked by age effects^20^. ^24^

**Figure 3:**
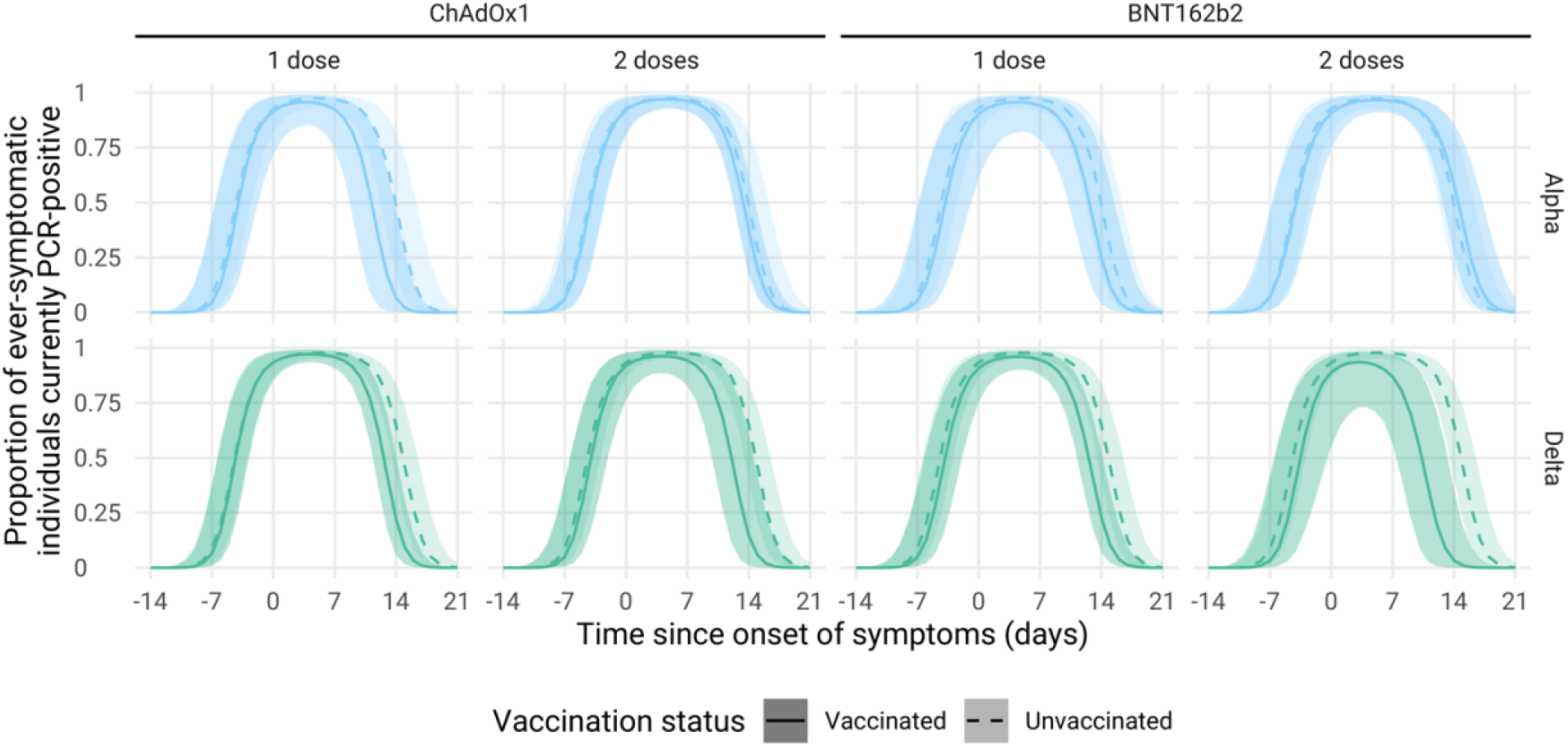
PCR positivity by Variant and vaccination status for symptomatic infections (index cases recruited from Pillar 2 testing and the symptomatic household contacts they infected). Lines represent median trajectories, and the ribbon is the 95% credible interval.

Our study comes with limitations, most importantly the potential for misclassification of the direction of transmission, the lack of inclusion of waning vaccine protection and the diversity of vaccines lineages and age groups in the dataset. To minimise the potential for misclassification we restrict the main analyses to only those putative transmission pairs where the there was no evidence against direct transmission based on phylogenetic distance (which was available for 63% of all putative transmission pairs) and where symptom onset in the contact did not pre-date that of the index case by more than 2 days. If residual misclassification between infector and infected remained this would re-attribute infection protection to transmission protection and vice versa. We also did not include waning of vaccine protection in our analyses^26^In the analysed dataset the longest reported time since vaccine receipt was 169 days. While some individuals in the analysis have since become eligible for booster vaccination over concerns of waning protection some of this potential effect will have been absorbed in our model in the age structuring because of the strong correlation between age and timing of vaccine eligibility as per vaccine roll-out strategy in the UK. Lastly, data collection spanned a period of multiple months during which Delta became the dominant strain in circulation in the UK and included participants vaccinated with two different vaccine products; thus requiring sub-strata analyses and reducing the effective sample size for each strata. We used a Bayesian model that allowed the borrowing of strength through the model hierarchy, and priors allowing us to make us of the heterogeneity in risk factors and not only estimate vaccine effectiveness against transmission in these strata but simultaneously estimate the difference in transmissibility in Alpha and Delta variants and the effectiveness of partially completed dosing schedules. The use of informative priors was integral to disentangling the confounded age and vaccine history effects which arose due to vaccine product prioritisation and were exacerbated by low counts for case-contact vaccine history combinations.

Our findings provide robust evidence from a prospective study that vaccination with either BNT162b2 or ChAdOx1 can help to substantially reduce, but not completely prevent, household transmission with SARS-CoV-2. This highlights the importance of vaccines to limit circulation of SARS-CoV-2 particularly in close and prolonged contact indoor settings. The effectiveness of booster doses to further enhance protection against transmission will need to be evaluated to better understand the extent to which we can rely on vaccination for the control of SARS-CoV-2 infection, particularly during winter seasons when most contacts occur in households or household-like settings.

## Data Availability

The data necessary to replicate results is available from the authors on request, subject to a data sharing agreement.

https://github.com/cmmid/hhSAR

## Authors’ contributions

EM developed the household transmission protocol; NA and JLB contributed to the study design; SF advised on the overall analytic approach; PW was responsible for developing and curating the database; CG, FK and CS assisted in data management; LL managed the team of study nurses; SC developed and conducted the Bayesian analysis; JH, SH and SC conducted the genomic analysis; SC, SF, EM and JH drafted the paper; SC and JH generated the tables and figures. All authors reviewed the manuscript prior to submission.

## Data sharing

All analysis code is available from https://github.com/cmmid/hhSAR. The data necessary to replicate results is available from the authors on request, subject to a data sharing agreement.

## Governance

The household surveillance protocol was approved by the UKHSA Research Ethics and Governance Group as part of the portfolio of the UKHSA’s enhanced surveillance activities in response to the pandemic. Oral informed consent for sampling and follow up was obtained by the nurses from household members who were free to decline to participate in the surveillance at any time. Consent for children was obtained by a parent or legal guardian. Only anonymised data were provided to non-UKHSA authors.

## Acknowledgements

We thank the nurses in the Immunisation Department of the UK Health Security Agency who recruited and followed up the households and the administrative staff who sent out the swabbing kits to households and arranged for their collection. We also thank the staff of the Virus Reference Department, Central Sequencing Laboratory and the Core Bioinformatics Team of PHE Colindale who performed the molecular testing and sequencing. Sequencing was financially supported in part by the COG-UK Consortium. COG-UK is supported by funding from the Medical Research Council (MRC) part of UK Research & Innovation (UKRI), the National Institute of Health Research (NIHR) and Genome Research Limited, operating as the Wellcome Sanger Institute. The authors also wish to thank Prof. Neil Ferguson (Imperial College London) and for his comments and questions about earlier versions of this analysis.

