## Appendix for "Effectiveness of BNT162b2 and ChAdOx1 against SARS-CoV-2 household transmission: a prospective cohort study in England"

#### Patient characteristics

Table A1: Number of contacts with listed vaccine status for each case vaccine status. Numbers in brackets show the additional individuals included in the sensitivity analysis.

| Case | 1 ChAdOx1 | 2 ChAdOx1 | 1 BNT162b2 | 2 BNT162b2 | None |
|---|---|---|---|---|---|
| 1 ChAdOx1 | 17 (4) | 1 | 3 | 4 | 23 (5) |
| 2 ChAdOx1 | 2 (1) | 26 (5) | 7 (1) | 12 (1) | 21 (9) |
| 1 BNT162b2 | 6 | 1 | 15 (2) | 2 | 33 (2) |
| 2 BNT162b2 | 9 | 8 | 4 | 10 | 9 |
| None | 6 | 2 | 4 | 5 | 48 (2) |

Cross-tabulation of contact vaccine status by household index case status is shown in Table A1, indicating that household contacts tend to have either the same vaccine status (product and number of doses) as their index cases or were not vaccinated at the time of data collection. Index cases who were unvaccinated rarely had household members who were vaccinated.

Table A2: Number of index cases and their household contacts with listed vaccine status for each age group. Numbers in brackets show the additional individuals included in the sensitivity analysis. There are no index cases younger than 18.

| Status | Vaccine | <18 | 18-49 | 50-64 | 65+ |
|---|---|---|---|---|---|
| Case | 1 ChAdOx1 | 0 | 15 (2) | 17 (2) | 3 (1) |
| | 2 ChAdOx1 | 0 | 20 (3) | 26 (4) | 1 |
| | 1 BNT162b2 | 0 | 22 (1) | 13 (2) | 2 |
| | 2 BNT162b2 | 0 | 13 | 17 | 1 |
| | None | 0 | 33 | 12 (1) | 0 |
| Contact | 1 ChAdOx1 | 0 (1) | 13 (2) | 22 (2) | 5 |
| | 2 ChAdOx1 | 0 | 13 (2) | 22 (2) | 3 (1) |
| | 1 BNT162b2 | 0 | 22 (2) | 9 | 2 (1) |
| | 2 BNT162b2 | 0 | 14 (1) | 16 | 3 |
| | None | 67 (7) | 55 (8) | 11 (2) | 1 (1) |

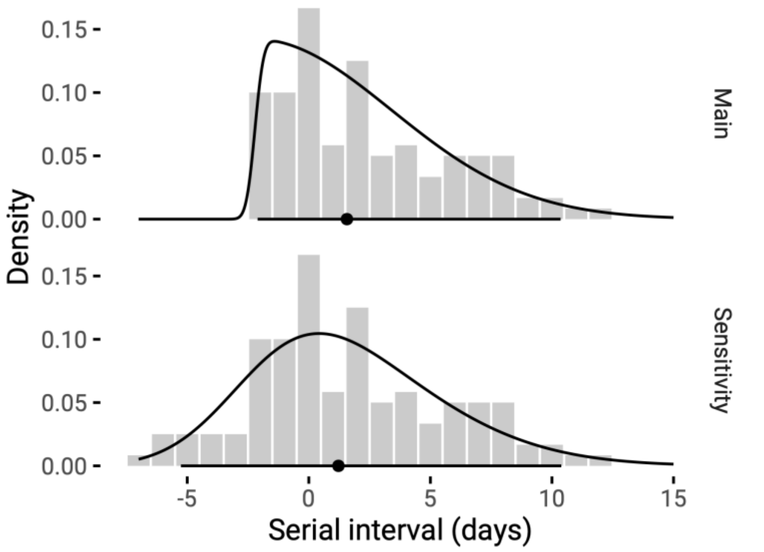


Figure A1: Distribution of contacts' observed serial intervals (grey bars). The black density curve represents the skewed normal distribution fit to the observed data, and its median and 95% range are shown as the black circle and line, respectively. The main analysis omits 16 households where any contact's serial interval is more negative than -2 days.

### Prevalence of lineages

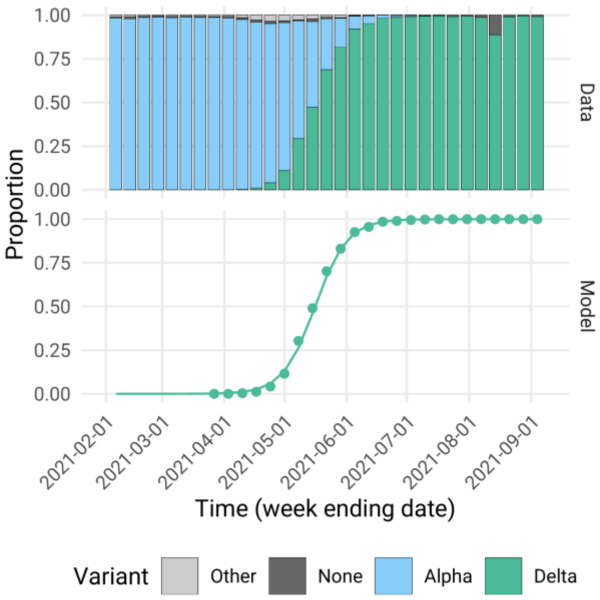


Figure A2: A) Weekly proportions of sequenced swabs from Pillar 2 testing that were Alpha, Delta, Other (known but not Alpha or Delta) or had no detected lineage. Only weeks after the commencement of the study are shown. B) Logistic regression model fit to the Alpha and Delta lineage swabs showing what proportion of these have Delta lineage (including AY sublineages of Delta).

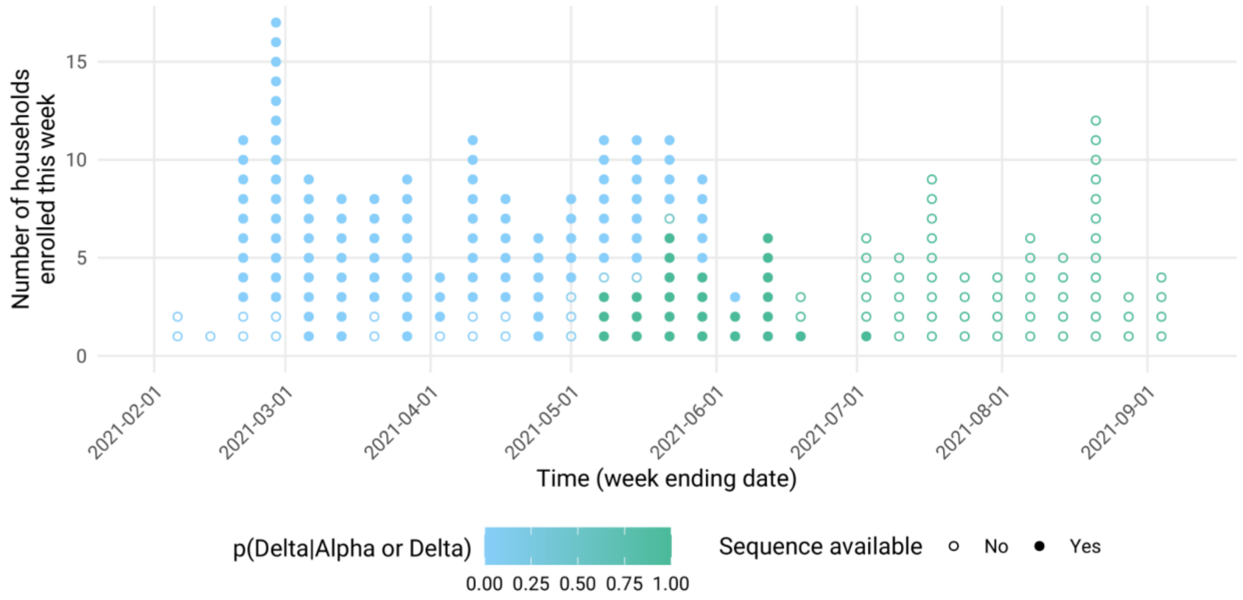


Figure A3: Lineage associated with each household, either through sequencing of study swabs (filled circles) or inferred through the model in Figure A2 (open circles). Inferred probability of Delta lineage is at time of the Day 1 swabs.

### Identification of non-household transmission

Whole-genome Illumina reads were retrieved from the European Nucleotide Database under the accession PRJEB37886. (https://www.ebi.ac.uk/ena/browser/view/PRJEB37886). Consensus genomes were generated using the Snippy pipeline mapping to the reference genome NC_045512.2.[1] Highly ambiguous and/or homoplasic sites were masked in the consensus alignment as described by de Maio et al.[2] A maximum-likelihood phylogeny was reconstructed from the consensus genomes under the Hasegawa-Kishino-Yano (HKY) model of nucleotide substitution with 1,000 ultrafast bootstrap replicates to assess branch supports and visualized in iTOL.[3,4]

ClusterPicker was used to identify clusters of transmission in the phylogeny.[5] These were defined as clusters of sequences with patristic distances of no more than 2 SNP ($6.6 \times 10^{-5}$ substitutions/site)[6] and bootstrap support of at least 70%.

### Model of Secondary Attack Rate

For the model of secondary attack rate the likelihood is

$$
\begin{aligned}
y_i &\sim \text{Bern}(p_i) \\
\log\left(\frac{p_i}{1-p_i}\right) &= \beta_0 + \delta_{V_i} + \\
&\quad \beta_{1,v_i,V_i}\mathbf{I}\left(d_{1,i,\text{contact}} \geq k_1\right) + \beta_{2,v_i,V_i}\mathbf{I}\left(d_{2,i,\text{contact}} \geq k_2\right) + \\
&\quad \gamma_{1,v_i,V_i}\mathbf{I}\left(d_{1,i,\text{case}} \geq k_1\right) + \gamma_{2,v_i,V_i}\mathbf{I}\left(d_{2,i,\text{case}} \geq k_2\right) + \\
&\quad \varepsilon_{\text{contact}}\mathbf{I}\left(A_{i,\text{contact}} < a_{\text{contact}}\right) + \varepsilon_{\text{case}}\mathbf{I}\left(A_{i,\text{case}} \geq a_{\text{case}}\right)
\end{aligned}
$$

where $\mathbf{I}$ is an indicator function which is 1 when its input is true and 0 otherwise and $d_{j,i,c}$ is the number of days since the $j$th dose of vaccine product $v$ was given to either contact $i$ or their household index case (indexed by $c$) who is infected with variant $V_i$. A fixed effect, $\delta$, accounts for the increased infectivity of Delta beyond that of Alpha. Protection afforded by dose $j$ is assumed to begin after $k = \{21,7\}$ days. These are currently fixed but a distribution may be used instead if there is some observed variability we wish to include. Age effects, $\varepsilon$, are assumed to be non-zero when the contact is younger than $A_{i,\text{contact}} = 18$ and when the case is at least as old as $A_{i,\text{case}} = 50$. Where $V_i$ was missing due to that household's swabs not being sequenced, it was sampled at each step of the MCMC from a Bernoulli distribution with its single parameter representing the modelled proportion of sequenced Pillar 2 swabs with Delta lineage at time of that household's Day 1 swabs.

The priors for the model parameters associated with transmission reduction are parameterised as weakly informative normal distributions (with means and precision ($\tau = \sigma^{-2}$))

$$
\begin{aligned}
\gamma_{j,v,V} &\sim \mathcal{N}\left(\gamma_{j,0,V}, \tau_\gamma\right) \\
\gamma_{j,0,V} &\sim \mathcal{N}\left(0, 10^6\right) \\
\tau_\gamma &= \sigma_\gamma^{-2} \\
\sigma_\gamma &\sim \text{Exp}\left(0.3\right)
\end{aligned}
$$

The penalised complexity prior on the standard deviation implies a prior probability of it exceeding 10 of 0.05.[24]

For the infection protection parameters, informative priors are derived from reported vaccine efficacy of the two vaccine products against the Alpha (B.1.1.7) and Delta (B.1.617.2) variants of SARS-CoV-2.[8] The priors are normally distributed for the log-odds ratios, with mean and precision parameters,

$$\begin{aligned}
\beta_{1,1,A} &\sim \mathcal{N}(\log 0.5, 2) & \beta_{1,1,D} &\sim \mathcal{N}(\log 0.65, 2)\\
\beta_{1,2,A} &\sim \mathcal{N}(\log 0.43, 0.25) & \beta_{1,2,D} &\sim \mathcal{N}(\log 0.67, 0.25)\\
\beta_{2,1,A} &\sim \mathcal{N}\left(\log \frac{0.25}{0.5}, 2\right) & \beta_{2,1,D} &\sim \mathcal{N}\left(\log \frac{0.33}{0.65}, 2\right)\\
\beta_{2,2,A} &\sim \mathcal{N}\left(\log \frac{0.06}{0.43}, 0.25\right) & \beta_{2,2,D} &\sim \mathcal{N}\left(\log \frac{0.12}{0.67}, 0.25\right)
\end{aligned}$$

Here we have taken the point estimates of the log odds ratios and scaled the standard errors up by a factor of 2, rounding to the nearest 0.5, in order to provide informative priors with additional variance that ensure that the posteriors are still sensitive to the data. As the $\beta_{2,v,V}$ represent the marginal effect of the second dose, we derive $\mathrm{B}_{2,v,V} = \beta_{1,v,V} + \beta_{2,v,V}$, the log-odds of the effect of double vaccination against variant $V$ with vaccine product $v$.

The effects of age have informative priors derived from Davies et al. for under-18s acquiring infection, $\varepsilon_{\text{contact}} \sim N(\log 0.50\,, 24)$, and from Yousaf et al.[16] cited in Goldstein et al.[11] for case transmission, $\varepsilon_{\text{case}} \sim N(\log 1.86\,, 4.67)$.

To determine how informative the priors above are, we replace the informative priors above for all $\beta_{j,v,V}$ with a weakly informative $N(0, \sigma_\beta^{-2})$ prior with $\sigma_\beta \sim \mathrm{Exp}(0.3)$ and the effects of age each having a weakly informative $N(0,10^{-6}\,)$ prior.

### Sensitivity analysis

Sensitivity analysis for the models described above was carried out by including all households, regardless of serial intervals. An additional 16 household index cases were included (all of which tested positive during the study) which contributed an additional 32 household contacts (of whom 23 tested positive). Sequences were available for mine of these additional index cases and ten of the contacts. Unless otherwise stated, the parameters given here represent transmission of a Delta infection from a non-elderly adult to an adult, neither of whom are vaccinated.

The additional infectiousness of Delta over Alpha was similar between the main (Risk Ratio: 1.64 (1.15, 2.44)), sensitivity analysis (1.58 (1.18, 2.21)), and model with weakly informative priors for all model effects (1.68 (1.17, 2.49)).

Age effects were comparable across the analyses (Table A3), The posteriors for the effect of age for child contacts in the main and sensitivity analyses is the same as the informative prior (to one decimal place) but even using the weakly informative prior the median and 2.5th percentile match those results. For 50+ year old adult index cases, the increase in infectivity is broadly similar for the main and sensitivity analyses, with the median similar to that of the informative prior used in its estimation. The posterior precision for the log-odds from the main and sensitivity analyses is approximately 17 and 19, respectively, compared to approximately 5 in the prior and 13 when using the weakly informative prior. So in the main analysis, the median may not have shifted much but there is more precision in the estimate.

Table A3: Posterior median risk ratios (and 95% CrIs) for effects of age, by analysis. *The Informative prior row is draws from the prior only.

| Analysis | Case 50+ | Contact <18 |
|---|---|---|
| Main | 1.1 (1.0, 1.2) | 0.8 (0.7, 1.0) |
| Sensitivity | 1.1 (1.0, 1.2) | 0.8 (0.7, 1.0) |
| Main with weakly informative priors | 1.1 (1.0, 1.2) | 0.9 (0.6, 1.0) |

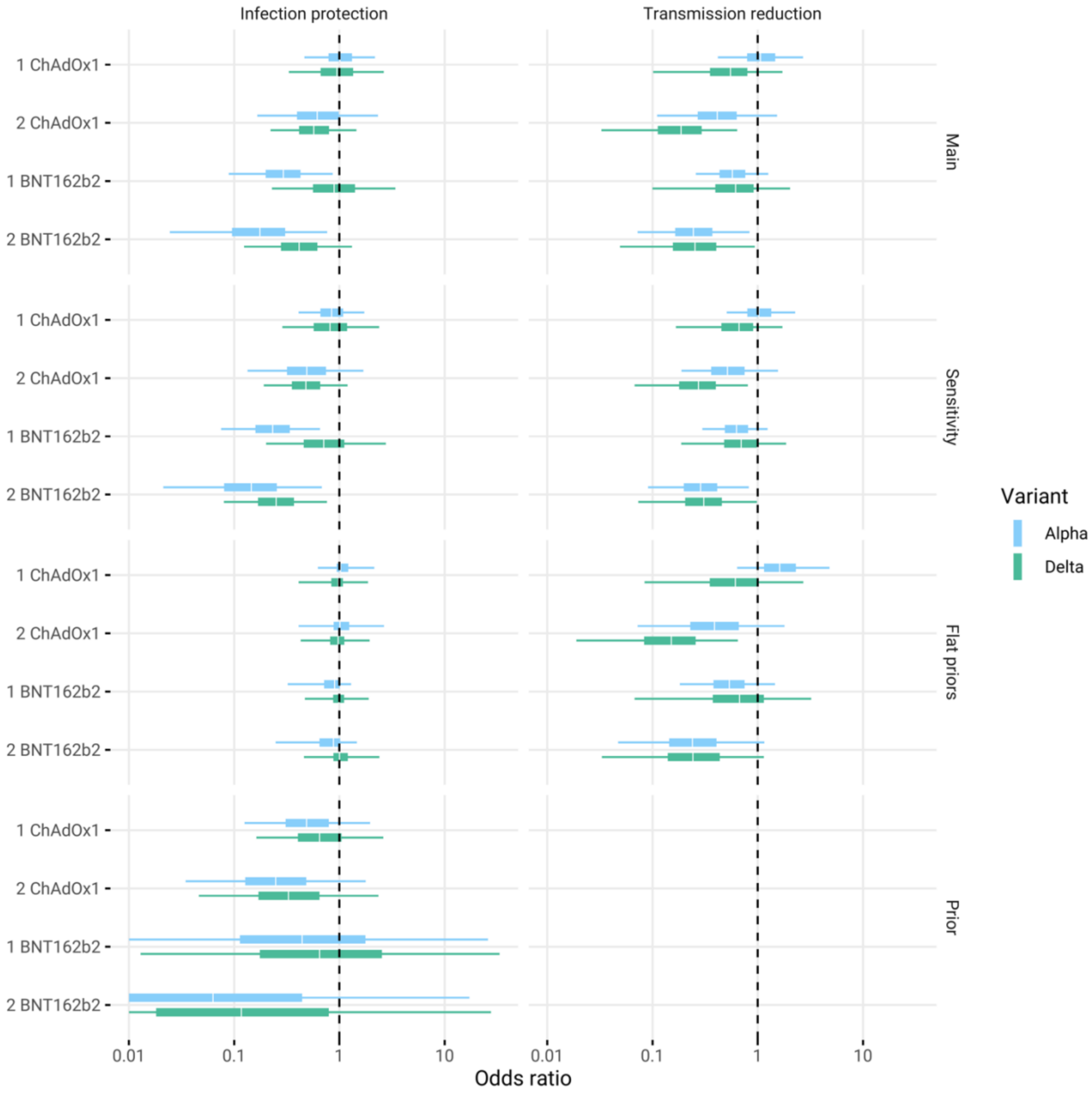


Figure A4: Odds Ratios (exponentiated regression coefficients) for effect of vaccination on transmission reduction in cases and protection against infection of contacts for main and sensitivity analysis (Sensitivity = all individuals, Flat priors = Main but with weakly informative priors), as well as ORs derived from sampling the priors (Priors). The 95% credible intervals for transmission reduction priors are $[0, \infty)$.

Table A4: Vaccine effectiveness against transmission reduction in cases and protection against infection of contacts for main and sensitivity analysis (Sensitivity = all individuals, Flat priors =

Main but with weakly informative priors. Presented as median and 95% credible interval.

| Analysis | Variant | Vaccine | Doses | Infection protection | Transmission reduction |
|---|---|---|---|---|---|
| Main | Alpha | ChAdOx1 | 1 | 3% (-38%, 39%) | -7% (-60%, 29%) |
| | | | 2 | 26% (-39%, 73%) | 35% (-26%, 74%) |
| | | BNT162b2 | 1 | 53% (7%, 83%) | 26% (-11%, 54%) |
| | | | 2 | 71% (12%, 95%) | 57% (5%, 85%) |
| | Delta | ChAdOx1 | 1 | 2% (-19%, 31%) | 14% (-11%, 52%) |
| | | | 2 | 14% (-5%, 46%) | 42% (14%, 69%) |
| | | BNT162b2 | 1 | 4% (-21%, 44%) | 9% (-16%, 49%) |
| | | | 2 | 24% (-2%, 64%) | 31% (-3%, 61%) |
| Sensitivity | Alpha | ChAdOx1 | 1 | 10% (-27%, 43%) | -4% (-55%, 28%) |
| | | | 2 | 36% (-23%, 76%) | 26% (-27%, 63%) |
| | | BNT162b2 | 1 | 61% (19%, 87%) | 21% (-20%, 50%) |
| | | | 2 | 74% (20%, 96%) | 51% (7%, 82%) |
| | Delta | ChAdOx1 | 1 | 4% (-16%, 34%) | 8% (-15%, 41%) |
| | | | 2 | 19% (-1%, 49%) | 30% (6%, 55%) |
| | | BNT162b2 | 1 | 9% (-14%, 49%) | 6% (-16%, 37%) |
| | | | 2 | 39% (7%, 72%) | 25% (-2%, 54%) |
| Flat priors | Alpha | ChAdOx1 | 1 | -5% (-49%, 22%) | -6% (-64%, 34%) |
| | | | 2 | -4% (-64%, 44%) | 44% (-22%, 78%) |
| | | BNT162b2 | 1 | 21% (-8%, 65%) | 29% (-10%, 58%) |
| | | | 2 | 27% (-11%, 79%) | 61% (10%, 87%) |
| | Delta | ChAdOx1 | 1 | 0% (-21%, 22%) | 16% (-12%, 53%) |
| | | | 2 | 2% (-14%, 28%) | 51% (24%, 73%) |
| | | BNT162b2 | 1 | 1% (-14%, 25%) | 13% (-14%, 55%) |
| | | | 2 | 5% (-10%, 39%) | 40% (8%, 65%) |

In the sensitivity analysis, no longer excluding households with extreme serial intervals, the only qualitative difference in conclusion, at 95% credibility, is that 2 doses of BNT162b2 protects contacts against infection against Delta lineage SARS-CoV-2 (the only vaccine status to do so) with a VE of 39% (95%: 7%, 72%) (Table A4). in the main analysis the median suggests a moderate effect but there is sufficient uncertainty in the estimate that it is difficult to distinguish from no effect (VE: 24%, 95%: -2%, 64%).

Table A5: Ratios of risk ratios for the effect of BNT162b2 vs ChAdOx1 for protection against infection of an adult contact (where case is unvaccinated) and transmission from a non-elderly adult cases to an unvaccinated adult contact.

| Variant | Dose | Infection protection | Transmission reduction |
|---|---|---|---|
| Alpha | 1 | 0.47 (0.16, 1.03) | 0.73 (0.41, 1.04) |
| | 2 | 0.40 (0.06, 1.70) | 0.74 (0.22, 1.64) |
| Delta | 1 | 0.97 (0.56, 1.41) | 1.02 (0.65, 1.69) |
| | 2 | 0.90 (0.49, 1.32) | 1.15 (0.80, 1.88) |

Sensitivity analysis indicates the same patterns of secondary attack rate by vaccination status hold for the default age group (Figure 2), with the exception of SARs that are approximately 0.1 (absolute value) higher for Delta households where the index case has received two doses of ChAdOx1 and the contact is either unvaccinated or has received at least one dose of ChAdOx1 (Figure A5). These differences still indicate that vaccinating index cases with two doses of ChAdOx1 is effective in reducing the SAR of Delta infections and the reduction for two doses is comparable in ChAdOx1 and BNT162b2. Figure A6 reflects that SARs are lower when the contact is a child and higher when the case is an adult over 50.

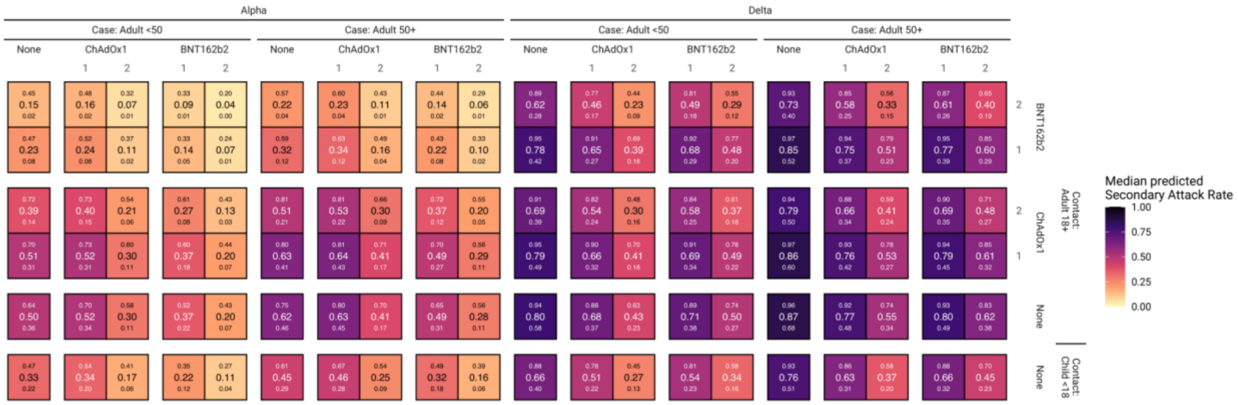


Figure A5: Main analysis shown in Figure 2, for each combination of index cases aged 18-49 and 50+ and contacts aged <18 and 18+. Large numbers inside cells are the median secondary attack rate (SAR), with the small numbers below and above corresponding to the 95% credible interval.

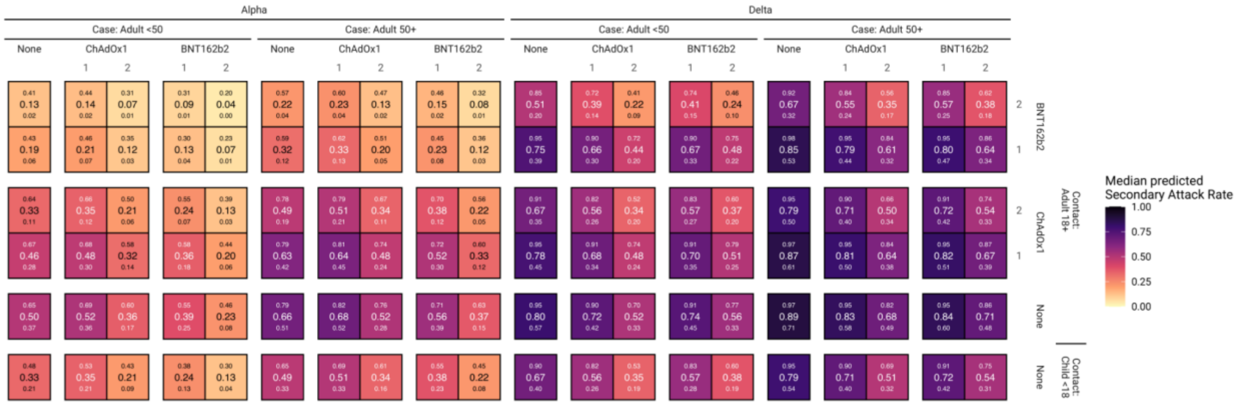


Figure A6: Sensitivity analysis (no households excluded for extreme serial intervals) of predicted secondary attack rates (as in Figure A5) for each combination of vaccine status of case and contact and each combination of index cases aged 18-49 and 50+ and contacts aged <18 and 18+.

### Infection history dynamics

We investigated PCR positivity after onset of symptoms for both cases and contacts by fitting a generalised linear mixed effects model with Bernoulli likelihood to each of the four vaccination groups. We assumed a quadratic effect for time since onset of symptoms, with random effect parameters by vaccine group (weakly informative priors with mean zero), to allow a rise from pre-exposure negativity, and a random intercept for each individual to account for their repeated measurement. Comparison was made against the no vaccination group.

The (up to) three swabs per individual were not part of routine surveillance with regular swabbing, and so negative swabs prior to the infecting exposure were unavailable. Therefore as part of the MCMC routine we sampled, at each step, an incubation period from $\Gamma(\alpha = 3.20, \beta = 0.56)$, being the distribution that matches a median of 5.1 days and 95th percentile of 11.7 days[12], and use this as a pseudo-absence (95% interval: 1.3, 13.4 days prior to onset of symptoms). Where the first test occurs prior to the onset of symptoms, the sampled incubation period distribution was truncated to the time of the first test to ensure that the infecting exposure occurred prior to both onset of symptoms and first positive test.

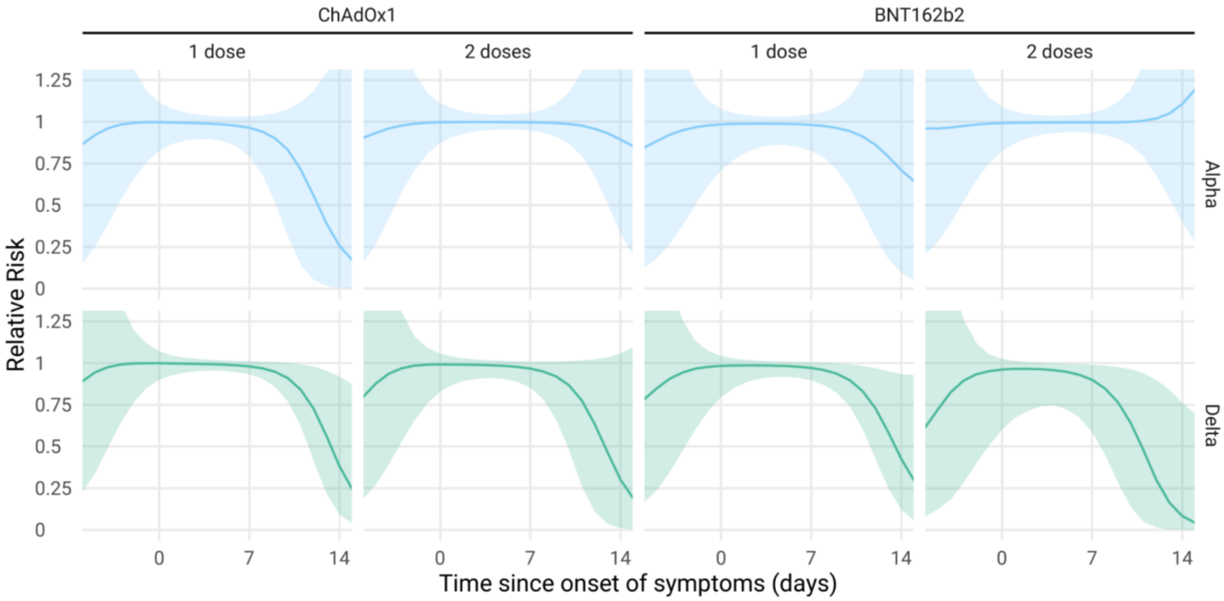


Figure A7: Relative risk of PCR positivity by Variant and vaccination status for symptomatic infections (index cases recruited from Pillar 2 testing and the symptomatic household contacts they infected). Lines represent median trajectories and the ribbon is the 95% credible interval.
